# Transportability of polysomnography-derived sleep physiological fingerprints across cohorts, measurement modalities and time: a four-axis evaluation in four cohorts

**DOI:** 10.64898/2026.11.21.26368655

**Authors:** Yulu Chen, Nai Ma, Qiao Deng, Xiaoming Lou, the Alzheimer’s Disease Neuroimaging Initiative

**Author notes:** Corresponding author: Qiao Deng, Department of Cerebrovascular Diseases, Ningbo No. 2 Hospital, Ningbo, China. Data used in preparation of this article were obtained from the Alzheimer’s Disease Neuroimaging Initiative (ADNI) database (adni.loni.usc.edu). As such, the investigators within the ADNI contributed to the design and implementation of ADNI and/or provided data but did not participate in analysis or writing of this report. A complete listing of ADNI investigators can be found at: http://adni.loni.usc.edu/wp-content/uploads/how_to_apply/ADNI_Acknowledgement_List.pdf.

## Abstract

Epidemiological evidence linking sleep to Alzheimer disease (AD) relies predominantly on self-reported sleep, whereas cohorts with polysomnography (PSG) and cohorts with AD biomarkers rarely share participants. Multifractal long-range-interdependency representations classify obstructive sleep apnoea severity accurately across cohorts, but whether such a physiological fingerprint can be transported between cohorts, between measurement modalities or over time has not been evaluated. We propose four axes of transportability and test them in the Sleep Heart Health Study (n = 5,614), MESA Sleep (n = 2,012), ADNI (n = 2,286) and MrOS Sleep (2,911 men; 1,026 re-examined after 6.37 years; 3,928 scored nights). Axis A1, cross-cohort construct convergence, passed: PSG-derived Sleep Coupling Disruption Index loadings agreed between SHHS and MESA (cosine 0.986; Wasserstein distance 0.0487). Axis A2, incremental validity beyond the apnoea-hypopnoea index, failed: the fully adjusted hazard ratio for incident cardiovascular disease was 1.009 (95% CI 0.941-1.085; p = 0.83) and the C-index was 0.8079 with or without the index; in MrOS, adding a multifractal coupling fingerprint increased R-squared by 0.00058 (likelihood-ratio p = 0.399). Axis A3, proxy identifiability, failed: the reliability ratio linking portable subjective complaints to objective physiological burden was 1.95 x 10-5, with a bootstrap 95% upper bound of 0.00261 and 0.0027 for a semantically optimised proxy, far below the 0.25 floor required for measurement-error correction. Under classical error this implies a 370-fold sample-size inflation; Monte-Carlo power at n = 10,000 was 0.177. A nominally significant association between night-time behaviour and dementia severity would require an implausible true physiological effect of 28.7 standard deviations, and it reversed to -0.0554 after adjustment for total neuropsychiatric burden. The prespecified ADNI primary outcome, plasma pTau217, was null (beta = 0.0033, p = 0.911), as were the MrOS six-year cognitive complaint outcome (beta = -0.0204, p = 0.375) and a 35-event self-reported dementia endpoint (OR = 0.921, p = 0.679). Axis A4, temporal stability, failed: six-year test-retest reliability was r = 0.157 (95% CI 0.096-0.216) for the fingerprint versus r = 0.455 (0.405-0.502) for AHI, despite stable concurrent validity (rho = 0.380 and 0.328 with same-night AHI). Spearman-Brown projection indicates that approximately nine nights must be averaged for the fingerprint to reach a reliability of 0.60, compared with two for AHI. A fingerprint can therefore be reproducible without being transportable. We provide a four-question decision workflow and an eight-item minimum reporting set; subjective sleep complaints should not be substituted for objective sleep physiology in aetiological inference.

## Introduction

Slow-wave sleep facilitates interstitial solute clearance [2,3], amyloid and tau deposition disturb sleep architecture [4], and cohort studies link sleep-disordered breathing and nocturnal hypoxaemia to mild cognitive impairment and dementia [5,6]. Loss of slow-wave sleep predicts incident dementia in community cohorts [7], and obstructive sleep apnoea has been related to longitudinal amyloid accumulation [8]. The AT(N) framework now permits these hypotheses to be tested against plasma and PET biomarkers in vivo [9,10]. The biological case for a sleep-AD relationship is therefore well established; the epidemiological difficulty is measurement.

Two structural problems recur. First, the large PSG cohorts that contain objective physiology - the Sleep Heart Health Study (SHHS) [11], MESA Sleep [12] and the National Sleep Research Resource more generally [13] - do not assemble cognitive or dementia outcomes, whereas AD cohorts such as ADNI [14] assemble plasma pTau217, amyloid and tau PET, MRI, cognitive composites and conversion endpoints but no PSG or actigraphy. No participant-level identifier is shared. Second, when a sleep variable is available in an AD cohort it is usually an informant-rated questionnaire item such as the night-time behaviour domain of the Neuropsychiatric Inventory (NPI-K) [15,16]. Subjective and objective sleep measures disagree systematically [17,18], so an association with a complaint item cannot be read as an effect of physiological sleep burden. The magnitude of the resulting exposure misclassification has rarely been quantified.

A third, newer problem concerns the representation of the physiological signal itself. Wang and colleagues [1] represented multichannel sleep signals as multifractal long-range-interdependency coupling matrices and classified apnoea severity with high accuracy across five cohorts, including SHHS and MESA. They identified extension beyond respiratory phenotyping as future work. Diagnostic accuracy, however, is a cross-sectional property. Before such a representation is used as a prognostic exposure, as a proxy in a cohort lacking PSG, or as a biomarker of a stable trait, its transportability must be established on at least four axes: convergence of the constructed phenotype across cohorts; incremental information beyond conventional indices; identifiability when only a proxy is measured; and stability over the intended prediction horizon. None of these is guaranteed by high classification accuracy.

We therefore executed a prespecified, real-data evaluation of these four axes across four cohorts. The programme produced predominantly null association results, which we report in full [31]. Its positive contribution is quantitative: we bound the reliability of subjective proxies for objective sleep burden, translate that bound into attenuation, sample-size and bias-re-estimation consequences, quantify the state-versus-trait behaviour of a multifractal coupling fingerprint over 6.4 years, and convert all of this into a decision workflow and a minimum reporting set for future sleep, cognition and signal-processing studies [30].

## Methods

### 2.1 Design and the four-axis framework

Four axes were defined before the synthesis (Fig 1B). A1, cross-cohort construct convergence, was considered satisfied when independently fitted component loadings agreed at cosine >= 0.90 and standardised score distributions had Wasserstein distance <= 0.10. A2, incremental validity, required a positive gain in discrimination or R-squared over a model already containing demographics, comorbidity and the apnoea-hypopnoea index (AHI), with likelihood-ratio p < 0.05. A3, proxy identifiability, required a reliability ratio lambda >= 0.25 for SIMEX or regression calibration [19,20]; 0.25 was the prespecified floor in the analysis plan and corresponds to an at most four-fold sample-size penalty. A4, temporal stability, required test-retest r >= 0.60 over the prediction horizon, the conventional threshold for group-level reliability [23]. Thresholds are pragmatic rather than universal; we report observed-to-threshold ratios for every axis so that readers may apply stricter criteria.

**Figure 1.**
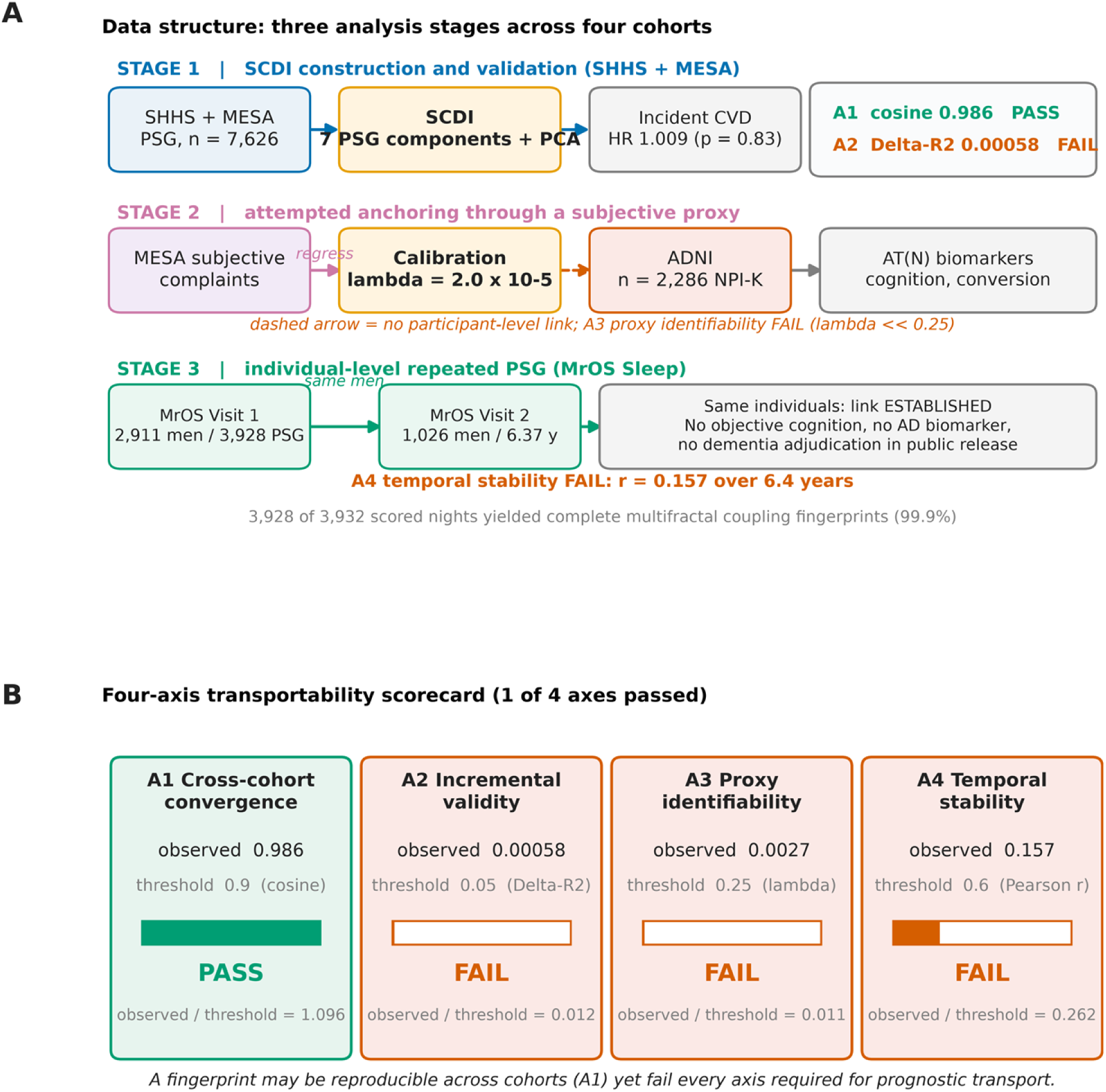
Study design and the four-axis transportability framework. (A) Data structure across three analysis stages and four cohorts. Stage 1 built a Sleep Coupling Disruption Index (SCDI) from seven PSG-derived components in SHHS (n = 5,614) and MESA Sleep (n = 2,012); Stage 2 attempted to calibrate a portable subjective-complaint proxy in MESA for transport into ADNI (n = 2,286 with Neuropsychiatric Inventory night-time behaviour, NPI-K); Stage 3 tested NPI-K against AT(N) biomarkers, cognition and conversion. NSRR sleep cohorts and ADNI share no participant-level identifier. The MrOS Sleep Study (n = 2,911 at Visit 1; 1,026 re-examined after 6.37 +/-0.98 years; 3,928 scored nights) provides an individual-level, repeated-PSG link, but its public release contains no objective cognitive test, AD biomarker or adjudicated dementia outcome. (B) Four axes required to transport a physiological fingerprint: A1 cross-cohort construct convergence, A2 incremental validity beyond the apnoea-hypopnoea index (AHI), A3 proxy identifiability (reliability ratio lambda), and A4 temporal stability. Only A1 was satisfied. Bars show the observed value relative to the prespecified threshold; cosine = 0.986, Delta-R2 = 0.00058, lambda = 0.0027 and Pearson r = 0.157.

### 2.2 Data sources and archive integrity

SHHS1 dataset 0.21.0 (5,804 participants), the SHHS CVD summary, SHHS1 5-min HRV series, SHHS1 EEG biomarkers and MESA Sleep dataset 0.4.0 (2,237 participants) were read in place from NSRR ZIP archives. ADNI tables (13 November 2026 export) comprised NPIQ, NPI, ADSP_PHC plasma/CSF/PET/T1/FLAIR/cognition, DXSUM, MMSE, CDR, ADAS, MoCA, GDS, PTDEMOG, APOERES, DESIKANLAB, VITALS, INITHEALTH, MUSE volumes, item-level data dictionaries and 1,526 502 x 502 resting-state connectivity matrices. MrOS Sleep 0.5.0 was supplied as six 200-GiB GNU tar split volumes totalling approximately 1 TB, one intermediate volume (part_af) unavailable at the time of analysis. All volumes were 512-byte-block aligned; part_aa began with the standard root header and part_ag ended with end-of-archive zero blocks. A virtual concatenated-stream reader sought tar headers without materialising the archive, recovering header chains by checksum-validated resynchronisation, and listed 11,315 members (7,864 annotation XML, 3,403 EDF, 12 CSV, 16 PDF, 2 MD). All 12 dataset CSV files and all 3,932 annotation XML files reside in intact volumes; the missing segment affects only raw EDF waveforms, which were not required here. Variable definitions were verified against the NSRR and ADNI dictionaries; several protocol assumptions were corrected during execution (MESA svspo25 is a hookup verification reading, not nocturnal hypoxaemia; self-reported sleep duration is stored in wkdaysleepdur5t; Centiloid values reside in ADSP_PHC_PET_Amyloid_Simple with -4 as not applicable; MESA Sleep 0.4.0 contains no BMI or CVD follow-up; MCI-to-AD conversion was reconstructed from longitudinal DXSUM because CONVERSIONS contains 19 records).

### 2.3 Stage 1: index construction, convergence and incremental validity

Seven PSG-derived components, each oriented so that larger values indicate more decompensation, were winsorised at the 1st/99th percentile, residualised on age, sex and race/ethnicity within cohort and standardised on SHHS moments: log1p(AHI), log1p(percent of total sleep time with SpO2 < 90%), 100 minus minimum SpO2, log1p(arousal index), log1p(WASO), 100 minus sleep efficiency, and log1p(|REM AHI - NREM AHI|) as a state-dependence term. A one-component PCA fitted in SHHS defined the Sleep Coupling Disruption Index (SCDI); the fit was repeated independently in MESA. Predictive validity used Cox models for incident CVD and CVD death in SHHS with censdate as time since baseline, adjustment for age, sex, race, BMI, smoking, hypertension, systolic pressure, diabetes and education, and exclusion of prevalent CVD. The protocol prespecified that absence of an AHI or CVD association would trigger a weight re-examination: no age adjustment; orthogonalisation of all components on log1p(AHI) followed by PCA of residuals, alone or blended with AHI; log1p(AHI) alone; and, as a post-hoc contrast, multifractal detrended fluctuation analysis [24,25] of 5-min cardiac interbeat series with detrended cross-correlation analysis [26] between HRV channels to obtain a long-range coupling fingerprint. The last analysis is explicitly exploratory (n = 310, 46 events).

### 2.4 Stage 2: calibration and the reliability ratio

MESA Sleep contains both subjective questionnaires and objective PSG and therefore served as the calibration set. The portable complaint score was the mean of four z-scored Women’s Health Initiative Insomnia Rating Scale items (difficulty falling asleep, frequent night waking, early awakening, difficulty returning to sleep). Four calibration models were fitted: the portable score; the first principal component of a 16-item subjective battery; physician-diagnosed insomnia alone; breathing symptoms alone; and a semantically optimised alignment that mapped MESA items onto the ADNI NPI-K sub-items NPIK1, NPIK2 and NPIK7. Ten-fold cross-validated R-squared and the partial R-squared attributable to the complaint were computed. The reliability ratio lambda = Var(E[SCDI | W]) / Var(SCDI), estimated as beta^2 Var(W) / Var(SCDI), is the quantity required for SIMEX and regression calibration. For the present synthesis we additionally obtained a nonparametric bootstrap distribution of lambda from 2,000 subject-level resamples of the calibration data.

### 2.5 Stage 3: ADNI association and specificity analyses

The exposure was baseline NPI-Q night-time behaviour burden (NPIKSEV when NPIK was endorsed, 0 otherwise), z-standardised; NPIK = 2, -1 and NPIKSEV = -4 were treated as missing codes. A dictionary audit identified richer phenotypes: NPI sub-items NPIK1, NPIK2, NPIK7 and NPIKTOT, clinician-rated insomnia in BLSCHECK/ADSXLIST, and insomnia history in INITHEALTH. NPI uses a skip pattern, so sub-items are unasked when the domain is not endorsed; treating these as missing produced a symptomatic-only subsample and implausible effect sizes (MMSE beta = 32). All refined analyses coded unendorsed domains as zero symptoms. Outcomes were harmonised plasma pTau217 (prespecified primary), pTau181, Abeta42, Abeta40, total tau, CSF Abeta42 and pTau181, Centiloid and amyloid status, tau PET meta-temporal SUVR, FLAIR WMH, SPARE-AD, hippocampal volume/ICV, PHC memory and executive composites, MMSE, CDR sum of boxes, ADAS-Cog 13 and MoCA. Cross-sectional models adjusted for age, sex, education, race and baseline diagnosis; extended models additionally adjusted APOE4 dose, polygenic hazard score, BMI and depressive symptoms. Benjamini-Hochberg FDR was applied within each outcome family. Linear mixed models estimated night-time burden x time interactions; Cox models estimated MCI-to-AD conversion. Specificity analyses added total NPI score; within-subject analyses demeaned exposure, outcome and visit month by participant and additionally adjusted visit-time trends and total NPI. Head motion (FD_postscrub) was a negative-control outcome. Measurement-error correction used an analytic lambda grid (0.05-0.75) and a Bayesian latent-variable sensitivity analysis with a Stage-2-informed prior.

### 2.6 Stage A: individual-level repeated PSG in MrOS

MrOS Sleep Visit 1 comprised 2,911 men (76.36 +/-5.47 years) with in-home PSG; Visit 2 re-examined 1,026 after 6.37 +/-0.98 years with repeat PSG. Each annotation file lists scored events with start time, duration and, for desaturations, SpO2 nadir and baseline. Events were mapped onto a common 30-s epoch axis to form five aligned channels: desaturation amplitude, respiratory-event count, arousal count, periodic-limb-movement count and scored sleep stage. MF-DFA [24,25] with second-order polynomial detrending over 8-10 log-spaced scales and q from -5 to +5 yielded the generalised Hurst exponent h(q), the q = 2 Hurst exponent and the multifractal spectral width; DCCA [26] between channels produced a 5 x 5 coupling matrix whose spectral radius, mean absolute off-diagonal coupling, maximum coupling and Fiedler value constituted the fingerprint. Event-interval series were supplementary. Completed features were obtained for 3,928 of 3,932 nights (99.9%). The prespecified primary model regressed Visit-2 sleep-related cognitive/functional interference on baseline z-standardised LRI spectral radius with HC3-robust ordinary least squares, adjusting for demographics, comorbidity, follow-up interval, baseline sleep complaint and conventional AHI. Twenty-five exposures were tested against nine outcome families with within-family FDR correction; incremental value was quantified by delta-R-squared and likelihood-ratio tests. Rare binary endpoints used Firth-penalised logistic regression [27] with standard errors from an explicit numerical Hessian of the penalised negative log-likelihood; a BFGS inverse-Hessian approximation had produced spuriously narrow intervals at 35 events and was discarded. Twelve sensitivity analyses, two negative-control outcomes, an attrition model, positive controls, history-consistency checks and cross-visit test-retest reliability were prespecified. The public MrOS release contains no objective cognitive test, AD biomarker or adjudicated dementia outcome; published MrOS analyses of cognitive decline [28,29] use data not distributed here.

### 2.7 Stage D: quantitative synthesis

Four derived quantities turned the null association results into transferable methodological statements. First, under classical measurement error with W = X + U, lambda = Var(X)/Var(W), the observed regression slope is attenuated to beta_obs = lambda x beta_true and the sample size required for a fixed power is inflated by 1/lambda. Required sample size was computed as n = (z_alpha/2 + z_beta)^2 / (beta_true^2 x lambda) for standardised variables, for beta_true in (0.10, 0.20, 0.30) and lambda in (0.0027, 0.0192, 0.05, 0.10, 0.25, 0.50, 0.75, 1.00). Second, bias re-estimation inverted the attenuation: an association reported for a proxy, forbidden to be interpreted as physiological, would require beta_true = beta_obs / lambda. We applied this to the observed CDR sum-of-boxes estimate and to hypothetical published standardised effects of 0.10-0.30. Third, Spearman-Brown projection, r_k = k r / (1 + (k - 1) r), gave the number of exchangeable nights that must be averaged for a measure with observed six-year reliability r to reach 0.60 or 0.80 [23]. Fourth, Monte-Carlo simulation validated the algebra: for each lambda and n (862, 2,000, 10,000), 600 replicates of X ∼ N(0,1), Y = 0.2 X + epsilon, W = X + U with Var(U) = (1 - lambda)/lambda were fitted by ordinary least squares; empirical power and the mean observed slope were recorded. Simulation output is reported only as a validity check and is labelled as such throughout.

### 2.8 Statistical analysis and reproducibility

Analyses used Python 3.13 (numpy, pandas, scipy, statsmodels, scikit-learn, lifelines, matplotlib, python-docx) with random seeds 20261116 for Stages 1-3 and 20261119 for Stages A and D. Complete cases were used; sample sizes are reported for every estimate. Fisher z transformation provided 95% confidence intervals for test-retest correlations. Reporting follows STROBE [30]. The study was designed so that a failure of any axis could be distinguished from an analysis artefact: positive controls, negative controls, concurrent validity, attrition models and history-consistency checks were prespecified.

### 2.9 Ethics and data availability

Data used in the preparation of this article were obtained from the Alzheimer’s Disease Neuroimaging Initiative (ADNI) database (adni.loni.usc.edu). ADNI was launched in 2003 as a public-private partnership led by Principal Investigator Michael W. Weiner, MD; its primary goal is to test whether serial magnetic resonance imaging, positron emission tomography, other biological markers and clinical and neuropsychological assessment can be combined to measure the progression of mild cognitive impairment and early Alzheimer disease. Sleep cohort data were obtained from the National Sleep Research Resource (https://sleepdata.org/) [13]. Data collection by ADNI and NSRR personnel is acknowledged in the Declarations.

All data were obtained under the respective NSRR and ADNI data-use agreements. Ethics approval and informed consent were obtained by the originating studies; the present analysis used de-identified secondary data only. The MrOS split archive was read in place and never modified. Source files are controlled-access and cannot be redistributed by the authors; the application routes, third-party contacts and the complete list of unrestricted derived data and analysis code are given in the Data Availability Statement under Declarations. Every number reported here can be regenerated by running the supplied code against source files obtained through those routes.

For human-participant reporting, this was a retrospective secondary analysis of archived cohort data, not a prospectively recruited study. Source files were first accessed for research purposes on 16 November 2026, and the final data extraction and all analyses were completed on 19 November 2026 (the ADNI tables were the 13 November 2026 export). The authors had no access to information that could identify individual participants at any time during or after data collection: every file released to the authors was de-identified and contained only coded study identifiers (SHHS, MESA and MrOS subject identifiers and ADNI RID/PTID), with no names, addresses, contact details or full dates of birth, and no linkage key was available. No participants were recruited for this analysis and no recruitment period therefore applies; no minors were enrolled in any of the four adult source cohorts. In the source studies all participants provided written informed consent before enrolment under approvals from the institutional review boards of the participating institutions; no consent waiver was sought or needed for the present analysis. Because the work used only de-identified third-party data and involved no new participant contact, no separate ethics approval was required for this analysis, which qualifies as exempt under the applicable IRB/ethics committee regulations. The work complied with the Belmont Report and the Declaration of Helsinki.

## Results

### 3.1 Cohorts and data integrity

Complete SCDI components were available for 5,614 SHHS and 2,012 MESA participants. The ADNI NPI-Q baseline sample comprised 2,286 participants (16.5% with any night-time behaviour disturbance), of whom 862 had plasma pTau217. MrOS contributed 2,906 Visit-1 and 1,022 Visit-2 nights with fingerprints and 1,021 men with two PSGs. All 12 dataset CSV and 3,932 annotation XML files were recovered from intact archive volumes (Table 1).

**Table 1.** Data assets and archive integrity across four cohorts.

| Cohort / release | N | Objective sleep | Cognitive / AD outcome | Role in this study |
|---|---|---|---|---|
| SHHS1 0.21.0 | 5,614 | PSG, AHI, hypoxaemia, HRV 5-min | None; adjudicated CVD | SCDI construction; A1; A2; CVD |
| MESA Sleep 0.4.0 | 2,012 | PSG plus sleep questionnaires | None | Independent SCDI fit; calibration (A3) |
| ADNI (13 Nov 2026) | 2,286 | None (NPI-K only) | pTau217, PET, MRI, cognition, conversion | Proxy-exposure associations; specificity |
| MrOS Sleep 0.5.0 | 2,911 / 1,026 | PSG twice, 3,928 scored nights | Self-reported interference and dementia only | Individual-level link; A4; LRI fingerprint |
| Archive integrity | 11,315 members | 12/12 CSV and 3,932/3,932 XML recovered | part_af absent at analysis; raw EDF only | Virtual tar seek; archives never modified |

### 3.2 A1: the construct replicates across cohorts

PCA loadings fitted independently in SHHS and MESA were nearly identical (cosine 0.986), explaining 39.8% and 43.2% of standardised variance; the standardised SCDI distributions had Wasserstein distance 0.0487 (Fig 2A). SCDI correlated strongly with AHI (rho 0.792 SHHS, 0.846 MESA) and hypoxic time (0.793, 0.871), moderately with arousal index (0.449, 0.477) and weakly with Epworth sleepiness (0.071, 0.067) (Fig 2B). SCDI tertiles were monotonically related to BMI (26.2, 27.7, 30.6 kg/m2) and AHI (4.4, 11.6, 27.7 events/h). A1 therefore passed.

**Figure 2.**
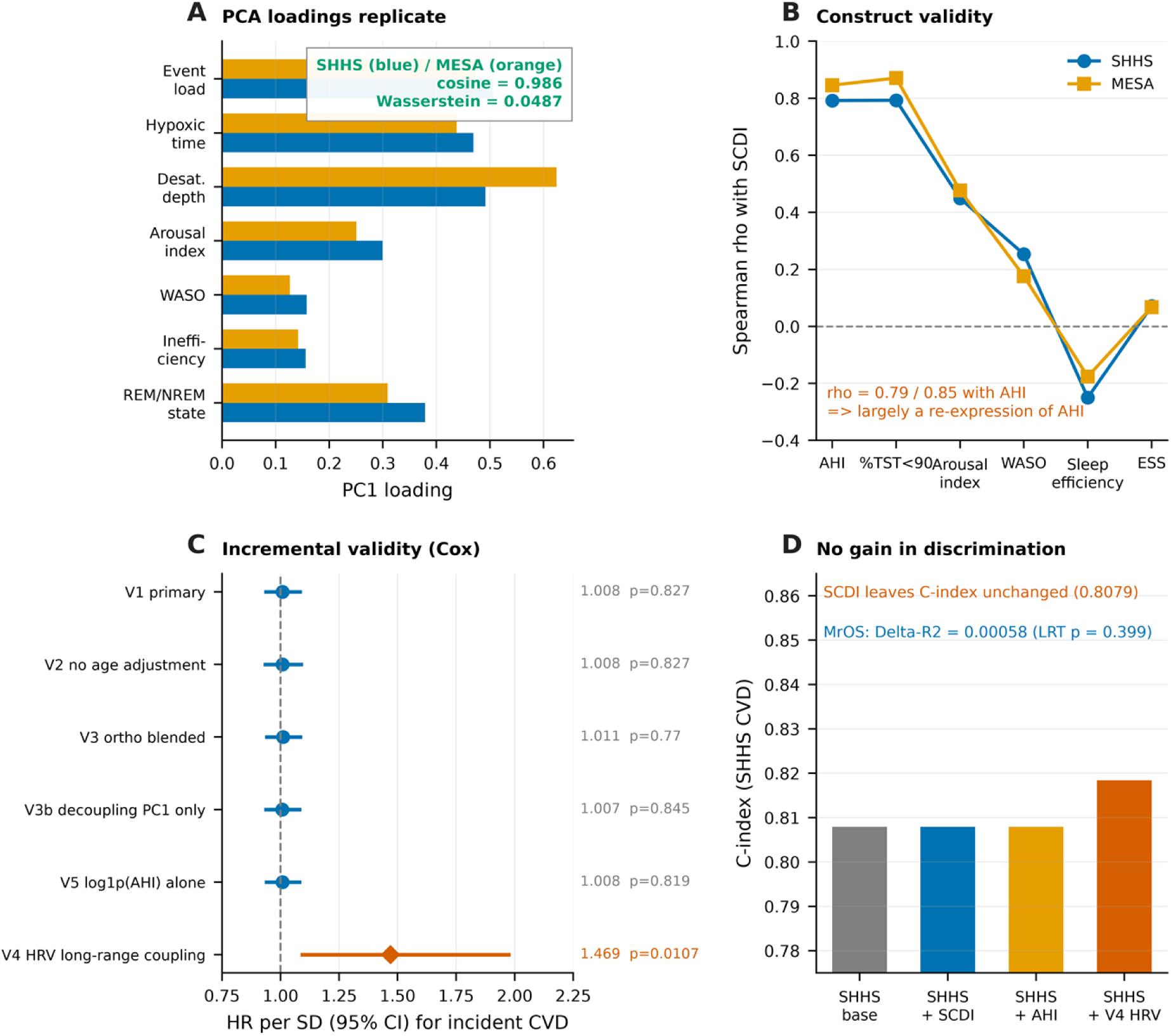
Reproducible construction without incremental value. (A) PC1 loadings of the seven SCDI components fitted independently in SHHS and MESA (cosine similarity 0.986; Wasserstein distance 0.0487 between standardised SCDI distributions). (B) Construct-validity correlations: SCDI correlates rho = 0.792/0.846 with AHI and 0.793/0.871 with percent of sleep time with SpO2 < 90%, but only 0.071/0.067 with Epworth sleepiness. (C) Fully adjusted Cox hazard ratios per SD for incident cardiovascular disease across five prespecified SCDI reweightings and the post-hoc HRV long-range coupling fingerprint (V4). Only V4 was significant (HR 1.466, 95% CI 1.095-1.964, p = 0.010; n = 310, 46 events; exploratory). (D) Discrimination: adding SCDI to a model containing demographics and AHI left the C-index unchanged at 0.8079; in MrOS, adding the LRI fingerprint increased R2 by 0.00058 (likelihood-ratio p = 0.399). Inset shows the MrOS R2 comparison on its true scale.

### 3.3 A2: reproducibility does not create incremental information

Among 3,776 SHHS participants without prevalent CVD and 754 incident events, the fully adjusted HR per SD of SCDI was 1.009 (95% CI 0.941-1.085; p = 0.83; C = 0.8079). Replacing SCDI with log1p(AHI), or adding both, did not change discrimination (C = 0.8079). All four prespecified reweightings and the pure AHI model produced HR between 1.007 and 1.011 with C between 0.8079 and 0.8099 (Fig 2C, D). The post-hoc HRV long-range coupling fingerprint was the only significant variant (HR 1.466, 95% CI 1.095-1.964; p = 0.010; bootstrap 95% CI 1.047-2.223; C 0.8049 to 0.8184; likelihood-ratio p = 0.0118), and was essentially orthogonal to AHI (rho = -0.037); it remains exploratory at 46 events. In MrOS, adding the LRI spectral radius to a model already containing demographics, comorbidity and AHI increased R-squared by 0.00058 (likelihood-ratio p = 0.399); mean coupling added 0.00237 (p = 0.088). A2 failed.

### 3.4 A3: the proxy is not identifiable, with quantitative consequences

Portable insomnia complaints did not predict SCDI (beta = +0.0044; p = 0.85; cross-validated R-squared = -0.0059; n = 1,995; Fig 3A). The bootstrap distribution of lambda had point estimate 1.95 x 10-5 and 95% CI upper bound 0.00261; the semantically optimised NPI-K-aligned model gave 0.0027 and the best subjective model of any kind gave 0.0192, all far below the 0.25 floor (Fig 3B). Under classical measurement error this implies attenuation to 0.27% of the true slope and a 370-fold sample-size inflation: for beta_true = 0.20 and 80% power, n rises from 196 to 72,674 (Fig 3C). Monte-Carlo simulation reproduced the expected slope (0.00057 observed versus 0.00054 theoretical at lambda = 0.0027) and showed that power reaches only 0.177 at n = 10,000, versus 1.000 when lambda = 1 (Fig 3D).

**Figure 3.**
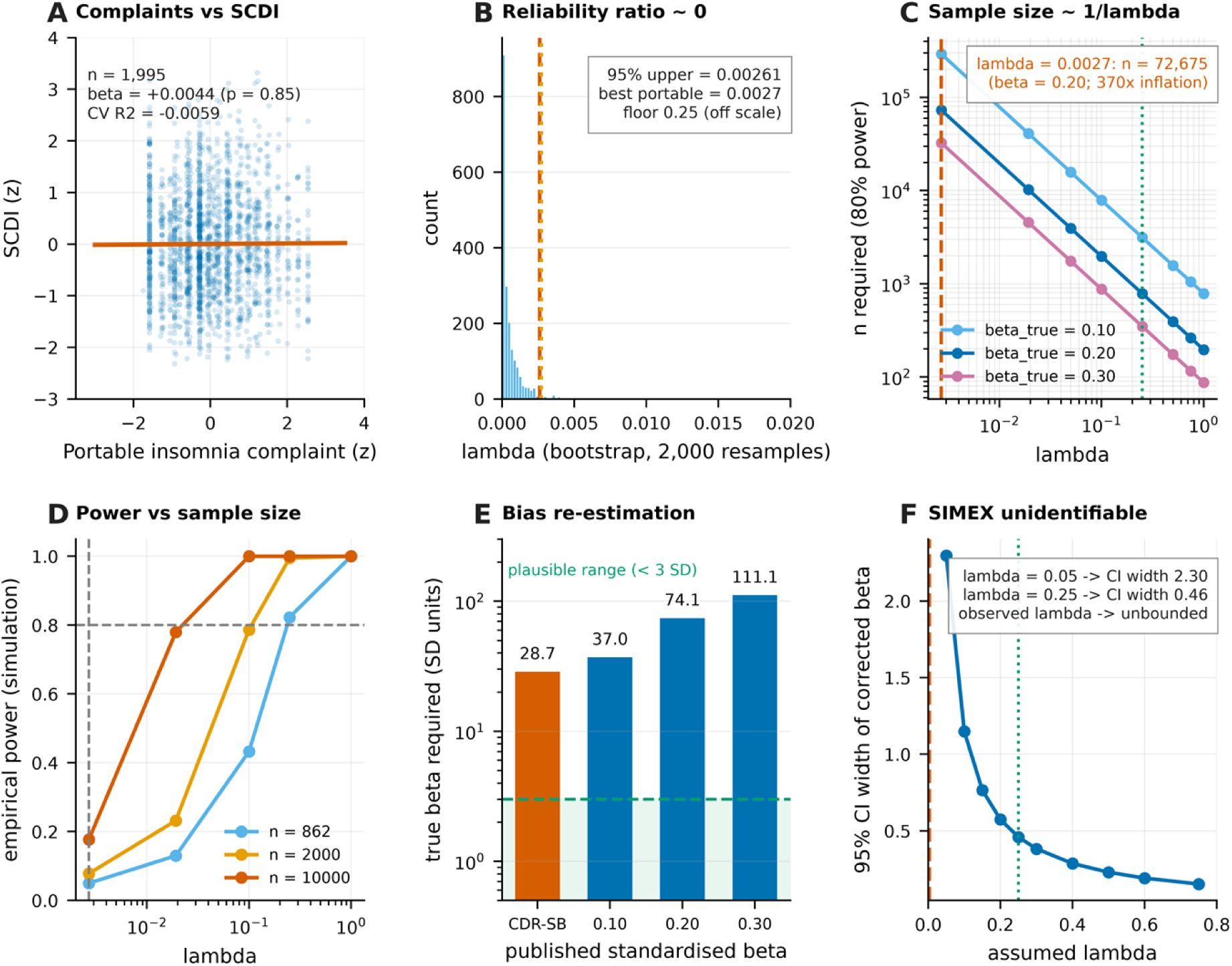
Quantitative identifiability and bias re-estimation. (A) Portable insomnia complaints versus objective SCDI in MESA (n = 1,995): beta = +0.0044 (p = 0.85), cross-validated R2 = -0.0059. (B) Nonparametric bootstrap of lambda (2,000 resamples): point estimate 1.95 x 10-5, 95% CI upper bound 0.00261; the semantically optimised NPI-K-aligned model gave 0.0027, both far below the prespecified SIMEX floor of 0.25. (C) Under classical measurement error, beta_obs = lambda x beta_true and the sample size required for 80% power is inflated by 1/lambda; at lambda = 0.0027 and beta_true = 0.20, n rises from 196 to 72,674. (D) Monte-Carlo validation (600 replicates per cell, beta_true = 0.20; simulation only): at the observed lambda, power remains 0.177 even with n = 10,000. (E) Re-estimation of published proxy associations: a nominally significant CDR sum-of-boxes association (beta = 0.0775) would require a true standardised physiological effect of 28.7 SD if it were driven by objective sleep burden; hypothetical published effects of 0.10-0.30 SD would require 37-111 SD. The shaded band marks the plausible range (< 3 SD). In ADNI the CDR-SB estimate reversed to -0.0554 after adjustment for total NPI burden. (F) Width of the 95% CI for the measurement-error-corrected pTau217 estimate as a function of assumed lambda; extrapolation to the observed lambda is unbounded.

Re-estimating published proxy associations gave implausible requirements (Fig 3E). The nominally significant CDR sum-of-boxes association (beta = 0.0775; p = 5.6 x 10-5) would require a true physiological effect of 28.7 SD at lambda = 0.0027, or 29.6 SD at the bootstrap upper bound; hypothetical published effects of 0.10-0.30 SD would require 37-111 SD. No value lies within the plausible range. Consistent with confounding rather than physiology, the CDR-SB estimate reversed to -0.0554 after adjustment for total NPI burden, and the strong within-person estimate (0.3093) fell to 0.2196 after controlling shared visit-time trends and to -0.0086 (p = 0.783) after additionally adjusting total NPI (Fig 4B). Analytic measurement-error correction was correspondingly uninformative: the corrected 95% CI widened from 0.46 at lambda = 0.25 to 2.30 at lambda = 0.05 and is unbounded at the observed lambda (Fig 3F). A3 failed.

**Figure 4.**
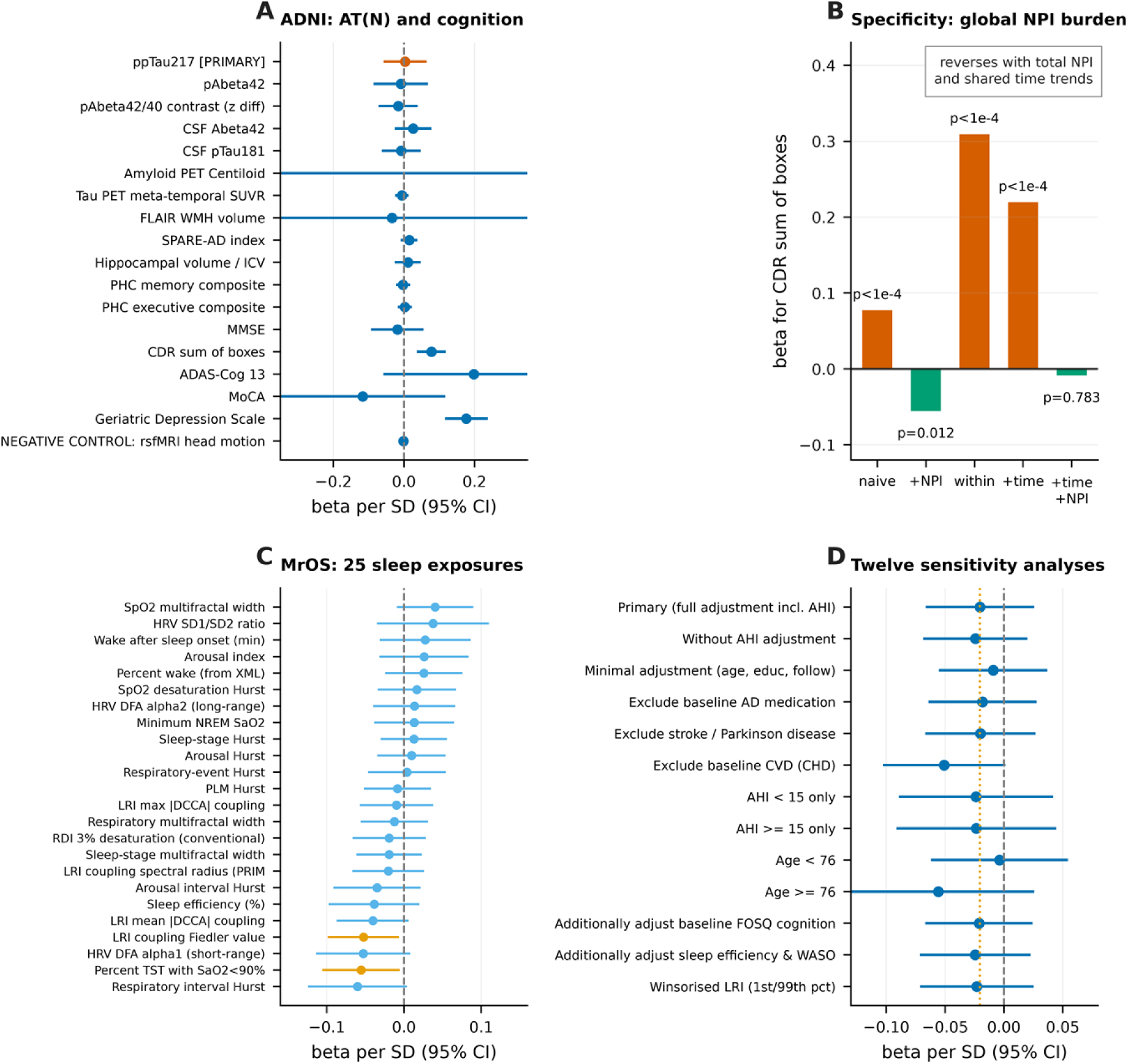
Association results in ADNI and MrOS. (A) Cross-sectional associations of baseline NPI-K night-time burden with 18 AT(N), imaging and cognitive outcomes in ADNI (beta per SD with 95% CI). The prespecified primary outcome, plasma pTau217, gave beta = 0.0033 (95% CI -0.0542 to 0.0608, p = 0.911, q = 0.911); no outcome survived Benjamini-Hochberg correction. (B) Specificity of the apparent CDR sum-of-boxes association: the naive between-person estimate (0.0775, p = 5.6 x 10-5) reversed after adjustment for total NPI score (−0.0554); the strong within-person estimate (0.3093) fell to 0.2196 after controlling shared visit-time trends and to -0.0086 (p = 0.783) after additionally adjusting total NPI. (C) All 25 baseline sleep exposures tested against the prespecified MrOS six-year cognitive/functional interference outcome; none survived false-discovery correction, and the two smallest nominal p-values were opposite in direction to the hypothesis. (D) Twelve sensitivity analyses of the MrOS primary estimate (range -0.0559 to -0.0038; all p > 0.05); the dotted line marks the primary estimate.

### 3.5 ADNI primary and secondary outcomes

Night-time behaviour was not associated with plasma pTau217 (beta = 0.0033 per SD; 95% CI -0.0542 to 0.0608; p = 0.911; q = 0.911; n = 862), nor with plasma pTau181, Abeta42, the Abeta42-Abeta40 contrast, CSF markers, Centiloid (beta = -1.027; p = 0.272), tau PET, WMH, SPARE-AD, hippocampal volume/ICV, cognitive composites, MMSE, MoCA or ADAS-Cog 13 (Fig 4A). Linear mixed models showed no exposure x time interaction for memory (p = 0.704; 11,990 observations) or executive function (p = 0.922). MCI-to-AD conversion gave HR 1.098 (0.979-1.232; p = 0.111). Of 55 tests on the refined sleep phenotypes, 19 reached nominal p < 0.05 but most pointed opposite to the hypothesis; the most consistent finding was an association with depressive symptoms, compatible with informant-reporting bias. Thirteen sensitivity and stratified analyses left the primary null estimate unchanged (S1 Fig).

### 3.6 A4: a valid state marker is a weak trait

In 1,021 men with two PSGs, LRI spectral radius correlated r = 0.157 (95% CI 0.096-0.216; Spearman rho = 0.139) across 6.37 years, whereas conventional RDI 3% correlated r = 0.455 (0.405-0.502; rho = 0.490) in 1,026 men (Fig 5A, B). Concurrent validity was stable at both visits (rho = 0.380 and 0.328 with same-night AHI), so the low longitudinal correlation is not a failure of the fingerprint to describe the night (Fig 5C). Spearman-Brown projection showed that reaching r >= 0.60 requires approximately nine nights for the LRI fingerprint and two for AHI; reaching 0.80 requires 22 and 5 nights (Fig 5D). The corresponding sample-size penalties for single-night measurement are 6.38-fold and 2.20-fold. A4 failed.

**Figure 5.**
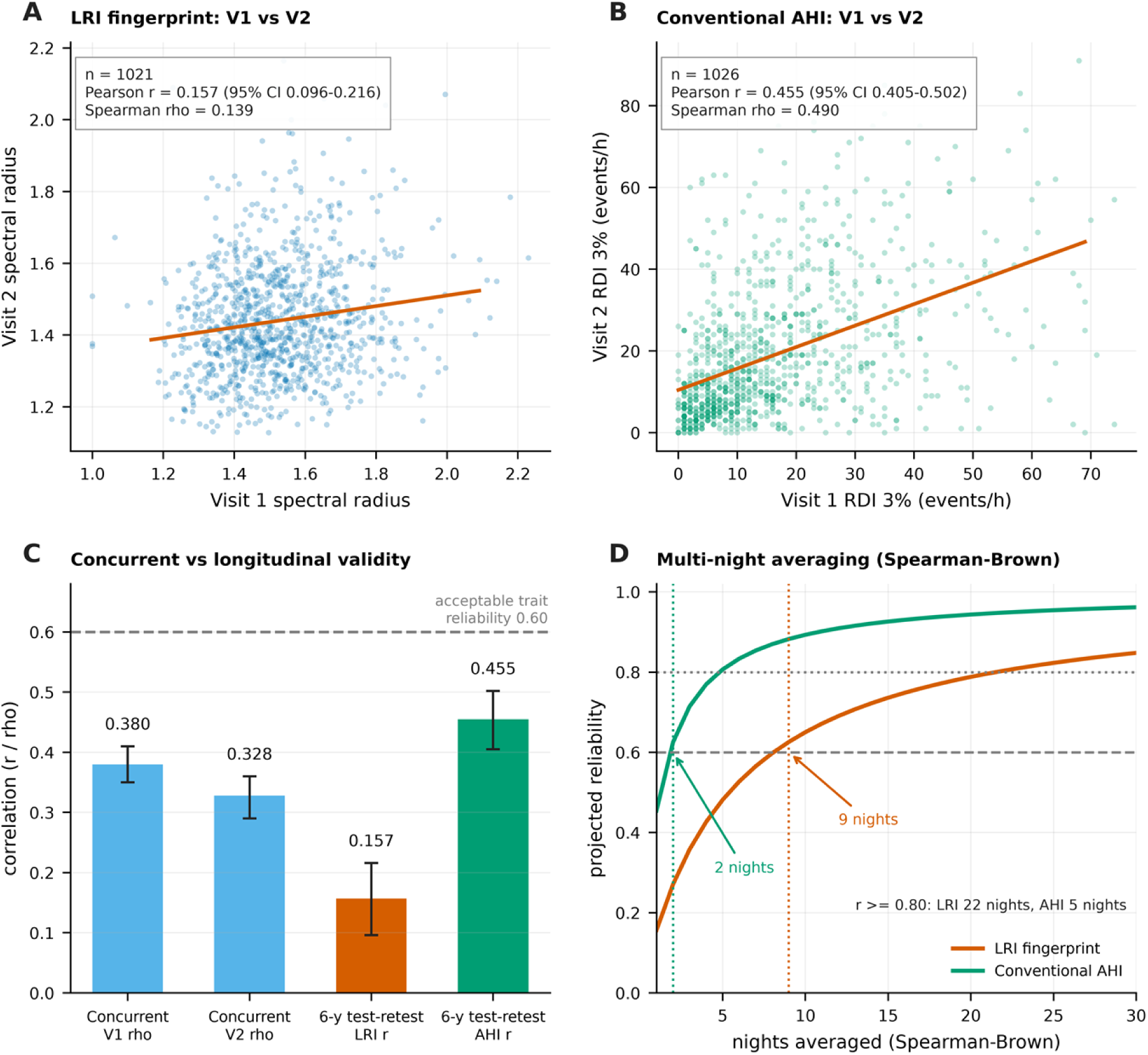
Temporal transportability: a valid state marker is a weak trait. (A, B) Visit-1 versus Visit-2 values of the LRI coupling spectral radius (n = 1,021; Pearson r = 0.157, 95% CI 0.096-0.216) and conventional RDI 3% (n = 1,026; r = 0.455, 95% CI 0.405-0.502) over 6.37 years. (C) The fingerprint retains concurrent validity with same-night AHI at both visits (Spearman rho = 0.380 and 0.328) but has far lower between-person stability than AHI. Error bars are Fisher-z 95% CIs; the dashed line indicates the conventional 0.60 reliability threshold. (D) Spearman-Brown projection of reliability as a function of the number of nights averaged: reaching r >= 0.60 requires approximately 9 nights for the LRI fingerprint and 2 nights for AHI (r >= 0.80 requires 22 and 5 nights respectively).

### 3.7 MrOS individual-level outcomes

The prespecified primary association was null (beta = -0.0204 per SD; 95% CI -0.0654 to 0.0247; p = 0.375; q = 0.643; n = 1,024; model R-squared = 0.1697). None of 25 exposures survived FDR correction; the two smallest nominal p-values (LRI Fiedler value p = 0.0217; percent of sleep time with SaO2 < 90% p = 0.0255; both q = 0.306) were opposite in direction to the hypothesis (Fig 4C). Twelve sensitivity analyses gave estimates from -0.0559 to -0.0038, all non-significant (Fig 4D). Among 35 self-reported dementia/AD events, no exposure was associated (LRI spectral radius OR = 0.921, 95% CI 0.625-1.358, p = 0.679; all p > 0.16 after excluding baseline AD-medication users; S1 Fig). A constructed incident cardiovascular endpoint was classified as indeterminate rather than a failed replication because positive controls were also null (age p = 0.789; diabetes p = 0.470; AHI p = 0.882) and 14.0-38.5% of baseline-positive lifetime histories reverted at follow-up (S1 Fig).

### 3.8 Scorecard

One of four axes passed (Fig 1B, Table 2). Observed-to-threshold ratios were 1.096 for A1, 0.012 for A2, 0.011 for A3 and 0.262 for A4. Because the axes are conjunctive - failure of any one invalidates the corresponding transported inference - the fingerprint can be constructed reproducibly but cannot be transported across measurement modalities or over a six-year horizon in these data.

**Table 2.** Four-axis transportability scorecard.

| Axis | Criterion | Observed | Observed / threshold | Verdict |
|---|---|---|---|---|
| A1 Cross-cohort convergence | cosine >= 0.90 | 0.986 (Wasserstein 0.0487) | 1.096 | PASS |
| A2 Incremental validity | delta-R2 > 0 and LRT p < 0.05 | 0.00058 (p = 0.399); C 0.8079 both | 0.012 | FAIL |
| A3 Proxy identifiability | lambda >= 0.25 | 0.0027 best proxy; 1.95 x 10-5 point | 0.011 | FAIL |
| A4 Temporal stability | test-retest r >= 0.60 | 0.157 (95% CI 0.096-0.216) | 0.262 | FAIL |

## Discussion

### 4.1 Principal findings

A PSG-derived physiological fingerprint can be reproduced almost exactly in two independent cohorts and still be useless for the inferences that motivate sleep and dementia research. In approximately 12,800 participants across four cohorts, the construct-convergence axis passed while incremental validity, proxy identifiability and temporal stability all failed. The results are internally coherent: an index that is largely a re-expression of AHI (rho = 0.79-0.85) cannot outperform AHI; a complaint item that explains essentially none of the variance in objective physiology cannot anchor two cohorts; and a descriptor whose six-year reliability is 0.157 cannot carry long-horizon prognostic information.

### 4.2 Reproducibility is not transportability

Cosine similarity of 0.986 between independently fitted loading vectors is a strong result and is worth reporting. It is also the least informative of the four axes, because the same linear combination of correlated respiratory events will emerge from any cohort with similar PSG scoring rules. High diagnostic accuracy in a cross-sectional task [1] is likewise compatible with failure on A2-A4. We therefore suggest that studies reporting a novel sleep phenotype separate construct replicability from transportability, and that the latter be treated as conjunctive: the observed-to-threshold ratios in Table 2 make explicit how far each axis falls short.

### 4.3 Quantitative re-estimation of the subjective-proxy literature

The most transferable result of this programme is a bound. Subjective insomnia complaints cannot recover objective PSG burden: lambda is at most 0.0027 even under the most favourable alignment, and its bootstrap upper bound is 0.00261. Under classical measurement error, a reported standardised effect of 0.10 with such a proxy cannot be a physiological effect - it would require a true effect of 37 SD. The observed CDR sum-of-boxes association in ADNI is a worked example: nominally strong (p = 5.6 x 10-5), it reversed after adjustment for total NPI burden and disappeared in within-person models once shared visit-time trends were accounted for. NPI-K is a component of the total NPI score and is rated by an informant whose assessment changes with disease severity. Studies that use NPI-K as a sleep variable should adjust for total NPI and depression and should distinguish informant-reported from self-reported symptoms. More generally, the discrepancy between subjective and objective sleep measures [17,18] should be treated as a quantified reliability problem rather than a qualitative caveat.

### 4.4 State markers, traits and the number of nights

The MrOS repeated-PSG analysis clarifies which part of the LRIA idea transfers. The multifractal coupling fingerprint retained concurrent validity with same-night AHI at both visits (rho = 0.380 and 0.328) but showed weak between-person stability over 6.4 years, whereas AHI was roughly three times more stable. Night-to-night variability in apnoea severity and physiological endotypes is well documented [21,22] and predicts clinical outcomes independently of a single-night AHI [22]; our result extends this from conventional indices to multifractal coupling features. The practical consequence is that a coupling fingerprint computed from one night should be described as a state marker. Using Spearman-Brown projection, roughly nine exchangeable nights are needed for this fingerprint to attain a reliability of 0.60, and 22 for 0.80, against two and five for AHI. Repeated-night averaging, not reweighting of derived indices, is the more defensible route to a trait-level exposure. The exploratory SHHS finding that an HRV long-range coupling fingerprint predicted incident CVD independently of AHI (HR 1.466, 46 events) is consistent with the view that long-range temporal structure carries information that event counts do not, but it requires external replication before it can be used to argue for prognostic value.

### 4.5 A decision workflow and reporting set

Fig 6A turns the four axes into four sequential questions. In these data the workflow stops at Q2: the fingerprint adds nothing beyond conventional severity, so even a reliable proxy or a stable trait would not rescue the inference. Table 5 and Fig 6B give the corresponding minimum reporting set. Three items are easy to omit and costly to omit. First, rare-event analyses must verify their standard-error implementation: an earlier run of our own pipeline used a BFGS inverse-Hessian approximation and produced an apparent p = 5.2 x 10-5 that became p = 0.733 with an explicit numerical Hessian. Second, self-reported lifetime disease history should be checked for reversals before defining incident endpoints; 14-38% of baseline-positive histories reverted in MrOS, and positive controls were null, so we classify that replication arm as indeterminate instead of negative. Third, informant-rated exposures should be adjusted for global symptom burden and shared time trends, as in the CDR-SB analysis. Reporting null results in full remains necessary for the evidence base [31].

**Figure 6.**
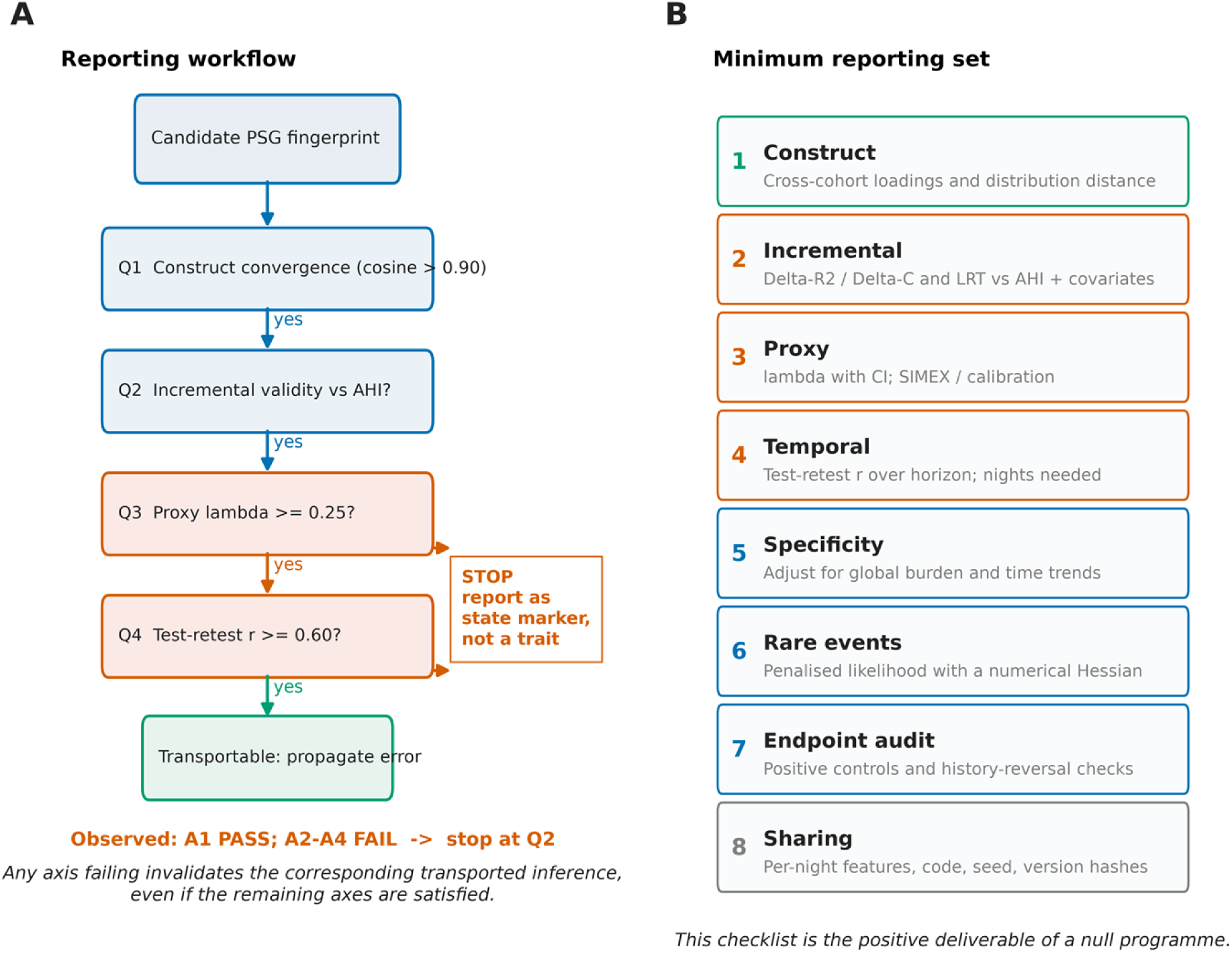
Proposed reporting workflow and minimum reporting set. (A) Four sequential questions to answer before a PSG-derived fingerprint is transported across cohorts, measurement modalities or time; failure at any axis invalidates the corresponding inference. In the present data only Q1 was satisfied. (B) Eight-item minimum reporting set covering construct convergence, incremental validity, proxy reliability, temporal stability, specificity, rare-event standard errors, endpoint audit and data/code sharing.

**Table 3.** Quantitative consequences of the observed reliability ratio.

| **Quantity** | **Value** | **Derivation / interpretation** |
|---|---|---|
| lambda point estimate | 1.95 x 10-5 | beta^2 Var(W)/Var(SCDI) in MESA, n = 1,995 |
| lambda bootstrap 95% upper | 0.00261 | 2,000 subject-level resamples |
| Best portable proxy lambda | 0.0027 | Semantically optimised NPI-K alignment |
| Slope attenuation | 0.27% | beta_obs = lambda x beta_true |
| Sample-size inflation | 370-fold | n = 196 -> 72,674 for beta_true = 0.20, 80% power |
| Monte-Carlo power at n = 10,000 | 0.177 | Simulation; beta_true = 0.20, 600 replicates |
| Required true beta for CDR-SB | 28.7 SD | 0.0775 / 0.0027; implausible (> 3 SD) |
| SIMEX 95% CI width | 0.46 at lambda = 0.25; 2.30 at 0.05 | Unbounded at observed lambda |
| Six-year reliability | r = 0.157 vs 0.455 (AHI) | Fisher-z 95% CIs 0.096-0.216 and 0.405-0.502 |
| Nights to r >= 0.60 | 9 (LRI) vs 2 (AHI) | Spearman-Brown; r >= 0.80 needs 22 vs 5 |

**Table 4.** Principal association estimates.

| Analysis | n / events | Estimate (95% CI) | p | q or LRT p |
|---|---|---|---|---|
| SHHS: SCDI -> incident CVD | 3,776 / 754 | HR 1.009 (0.941-1.085) | 0.83 | C = 0.8079 |
| SHHS: HRV coupling -> CVD (exploratory) | 310 / 46 | HR 1.466 (1.095-1.964) | 0.010 | LRT 0.0118 |
| ADNI: NPI-K -> plasma pTau217 (primary) | 862 | beta 0.0033 (-0.0542-0.0608) | 0.911 | q = 0.911 |
| ADNI: NPI-K -> CDR-SB, naive | 2,263 | beta 0.0775 | 5.6 x 10-5 | reverses to -0.0554 |
| ADNI: NPI-K -> CDR-SB, within-person + time + NPI | 3,306 obs / 788 | beta -0.0086 | 0.783 | - |
| ADNI: MCI -> AD conversion | 550 / 247 | HR 1.098 (0.979-1.232) | 0.111 | C = 0.644 |
| MrOS: LRI -> 6-y cognitive interference (primary) | 1,024 | beta -0.0204 (-0.0654-0.0247) | 0.375 | q = 0.643 |
| MrOS: incremental delta-R2 beyond AHI | 1,024 | 0.00058 | 0.399 | LRT 0.399 |
| MrOS: LRI -> self-reported dementia/AD | 1,025 / 35 | OR 0.921 (0.625-1.358) | 0.679 | Firth |

**Table 5.** Minimum reporting set for studies transporting a sleep fingerprint.

| # | **Item** | **What must be reported** |
|---|---|---|
| 1 | Construct | Cross-cohort loading similarity and distribution distance |
| 2 | Incremental | delta-R2 / delta-C and a likelihood-ratio test against AHI plus covariates |
| 3 | Proxy | Reliability ratio lambda with CI; SIMEX or regression calibration |
| 4 | Temporal | Test-retest r over the prediction horizon; nights required |
| 5 | Specificity | Adjustment for global symptom burden and shared time trends |
| 6 | Rare events | Penalised likelihood with an explicit numerical Hessian |
| 7 | Endpoint audit | Positive controls and history-reversal checks |
| 8 | Sharing | Per-night features, code, random seed and version hashes |

### 4.6 Limitations

The public MrOS Sleep release contains no objective cognitive test, AD biomarker, dementia adjudication, dated events or mortality follow-up; all cognitive endpoints used here are self-reported and the dementia endpoint has 35 events. This study therefore cannot determine whether objectively measured sleep physiology predicts AD pathology, and our null MrOS results should not be read as evidence against the sleep-AD hypothesis. MrOS cognitive data exist [28,29] and should be obtained from the coordinating centre; the 3,928 per-night features and all merging code can then be reused. ADNI has no PSG or actigraphy, and plasma/PET measurements were not always contemporaneous with the NPI assessment (244 identical-visit pairs). Population differences between community-based MESA/SHHS and the largely non-Hispanic White ADNI clinical cohort limit comparability. We did not recompute the full LRIA representation from raw waveforms; the fingerprint here is derived from scored event annotations, an intentional and reproducible simplification. The derivation of sample-size inflation assumes classical, non-differential measurement error and a single standardised exposure; differential error, as is likely with informant reporting, may bias in either direction and our 370-fold figure should be read as a lower bound on the problem rather than an exact correction. Complete-case analysis was used; multiple imputation, mediation and competing-risk models were not performed because primary effects were null. The HRV result is exploratory and based on 46 events. Finally, our thresholds (cosine 0.90, lambda 0.25, r 0.60) are pragmatic; the observed-to-threshold ratios are reported so that the framework can be re-evaluated under stricter criteria.

## Conclusions

Reproducibility is not transportability. A PSG-derived multifractal coupling fingerprint replicated almost identically in two independent cohorts (cosine similarity 0.986) and carried information within a single night, yet failed the three tests that would justify moving it into an Alzheimer disease cohort: it added no prognostic value beyond AHI (Cox HR 1.009, p = 0.83; C-index 0.8079 for both models), could not be reconstructed from subjective complaints, and was only weakly stable over 6.4 years (r = 0.157 versus 0.455 for AHI). One of four axes passed. These results do not argue against a sleep-AD relationship; they delimit what can be inferred from these four cohorts. With the exposures, proxies and follow-up available here, the phenotype is not identifiable across cohorts, modalities or long prediction horizons, and an apparently strong association with a subjective sleep item is more parsimoniously explained by shared neuropsychiatric burden and visit-time trends than by sleep physiology.

The transferable output is a set of quantitative boundaries rather than a positive association. Three are hard enough to serve as design constraints. First, the reliability ratio linking subjective complaints to objective PSG burden stayed below 0.003 for any portable measure (point estimate 1.95 x 10-5; bootstrap upper bound 0.00261), which inflates the sample size needed to detect a standardised effect of 0.20 from 196 to 72,674 - a 370-fold penalty. Second, re-expressing the naive CDR-SB association as a physiological effect would require a true effect of 28.7 SD, outside the plausible range. Third, a single night of the coupling index behaves as a state rather than a trait, requiring nine nights of averaging to reach r = 0.60 and 22 nights to reach 0.80. Each bound is reported with its observed-to-threshold ratio in Table 2 so that it can be recomputed under different assumptions.

We therefore propose the four-axis scorecard as a minimum reporting set for studies that transport a sleep phenotype to a new outcome, cohort or measurement modality, and as a checklist at the design stage: establish incremental validity over conventional severity before invoking a novel marker, quantify the reliability ratio before using a questionnaire as a proxy for physiology, and estimate temporal stability before treating one night as a trait. A decisive test of the sleep-AD hypothesis requires a cohort in which repeated objective sleep measurement, adjudicated cognitive trajectories and AD biomarkers are collected in the same individuals. Until such data exist, subjective sleep items, objective PSG physiology and neurodegeneration should not be used as interchangeable exposures.

## Declarations

### Ethics approval and consent to participate

the Sleep Heart Health Study, the Multi-Ethnic Study of Atherosclerosis, the MrOS Sleep Study and the Alzheimer’s Disease Neuroimaging Initiative were approved by the institutional review boards of their participating institutions, and all participants provided written informed consent before enrolment. The present work is a secondary analysis of de-identified data released under the NSRR Data Access and Use Agreement and the ADNI Data Use Agreement. No new participants were recruited, no identifying information was accessed and no additional contact with participants took place; consequently no separate ethical approval was required for this analysis. All procedures were conducted in accordance with the Declaration of Helsinki and with the data-use and publication policies of the source studies.

## Consent for publication

not applicable.

## Availability of data and materials

all source data are third-party, controlled-access data. The authors had no special access privileges and obtained them through the same application routes open to any qualified investigator; no author-mediated data request is needed. Sleep cohort data (Sleep Heart Health Study, MESA Sleep and MrOS Sleep) are distributed by the National Sleep Research Resource at https://sleepdata.org/. Access requires a free account, a completed Data Access and Use Agreement describing the proposed use, and approval by the NSRR Review Committee; enquiries should be directed to (see https://sleepdata.org/about/data-security). ADNI data (13 November 2026 export) are distributed through the LONI Image and Data Archive at https://adni.loni.usc.edu/; access requires acceptance of the ADNI Data Use Agreement and an online application reviewed by the ADNI Data Sharing and Publications Committee (enquiries). Both agreements prohibit redistribution of source files by data users, which is why raw EDF waveforms, XML event annotations and individual-level ADNI tables cannot be deposited publicly by the authors. These restrictions apply to source data only and do not apply to any derived product reported here. Requests for source data should be directed to the repositories and committees named above rather than to the authors.

The minimal data set underlying every figure, table and numerical result is unrestricted and is submitted as Supporting Information: the per-night multifractal coupling feature matrix (mros_xml_features.csv, 3,928 rows x 53 columns); the Stage D result set (stageD_results.json and six CSV tables covering the lambda bootstrap, power-versus-lambda, bias re-estimation, Spearman-Brown projections, Monte-Carlo attenuation and the four-axis scorecard); the manuscript validation log and the field-level reference audit log; the complete analysis code (virtual tar reader, MF-DFA/DCCA, index construction, calibration, ADNI and MrOS association models, Firth module, quantitative synthesis and figure generation); and all figure files in PNG, PDF and SVG format. Together with source data obtained through the routes above, these files reproduce every table and figure in this article. The MrOS part_af volume was unavailable at the time of analysis and contains raw EDF waveforms only; all 12 dataset CSV files and all 3,932 annotation XML files used here reside in intact volumes, so no derived feature depends on that segment. Derived products are made available under the Creative Commons Attribution 4.0 International licence (CC BY 4.0).

## Competing interests

the authors declare that they have no competing interests.

## Funding

this work received no specific grant from any funding agency in the public, commercial or not-for-profit sectors. Data collection and sharing for the source cohorts was funded separately, as acknowledged below.

## Authors’ contributions

Yulu Chen: data curation, validation, writing - review and editing. Nai Ma: data curation, validation, writing - review and editing. Qiao Deng: conceptualization, data curation, formal analysis, methodology, software, visualization, writing - original draft. Xiaoming Lou: funding acquisition, project administration, supervision, writing - review and editing. Qiao Deng and Xiaoming Lou contributed equally to this work. All authors read and approved the final manuscript.

## Data Availability

all source data are third-party, controlled-access data. The authors had no special access privileges and obtained them through the same application routes open to any qualified investigator no author-mediated data request is needed. Sleep cohort data (Sleep Heart Health Study, MESA Sleep and MrOS Sleep) are distributed by the National Sleep Research Resource at https://sleepdata.org/. Access requires a free account, a completed Data Access and Use Agreement describing the proposed use, and approval by the NSRR Review Committee enquiries should be directed to (see https://sleepdata.org/about/data-security). ADNI data (13 November 2026 export) are distributed through the LONI Image and Data Archive at https://adni.loni.usc.edu/ access requires acceptance of the ADNI Data Use Agreement and an online application reviewed by the ADNI Data Sharing and Publications Committee (enquiries). Both agreements prohibit redistribution of source files by data users, which is why raw EDF waveforms, XML event annotations and individual-level ADNI tables cannot be deposited publicly by the authors. These restrictions apply to source data only and do not apply to any derived product reported here. Requests for source data should be directed to the repositories and committees named above rather than to the authors. The minimal data set underlying every figure, table and numerical result is unrestricted and is submitted as Supporting Information: the per-night multifractal coupling feature matrix (mros_xml_features.csv, 3,928 rows x 53 columns) the Stage D result set (stageD_results.json and six CSV tables covering the lambda bootstrap, power-versus-lambda, bias re-estimation, Spearman-Brown projections, Monte-Carlo attenuation and the four-axis scorecard) the manuscript validation log and the field-level reference audit log the complete analysis code (virtual tar reader, MF-DFA/DCCA, index construction, calibration, ADNI and MrOS association models, Firth module, quantitative synthesis and figure generation) and all figure files in PNG, PDF and SVG format. Together with source data obtained through the routes above, these files reproduce every table and figure in this article. The missing MrOS part_af volume affects raw EDF waveforms only all 12 dataset CSV files and all 3,932 annotation XML files used here reside in intact volumes, so no derived feature depends on the absent segment. Derived products are made available under the Creative Commons Attribution 4.0 International licence (CC BY 4.0).

## Acknowledgements

the authors thank the investigators, staff and participants of the Sleep Heart Health Study, the Multi-Ethnic Study of Atherosclerosis, the MrOS Sleep Study and the Alzheimer’s Disease Neuroimaging Initiative, and the National Sleep Research Resource for making these resources available. Data collection and sharing for this project was funded by the Alzheimer’s Disease Neuroimaging Initiative (ADNI) (National Institutes of Health Grant U01 AG024904) and DOD ADNI (Department of Defense award number W81XWH-12-2-0012). ADNI is funded by the National Institute on Aging, the National Institute of Biomedical Imaging and Bioengineering, and through generous contributions from the following: AbbVie; Alzheimer’s Association; Alzheimer’s Drug Discovery Foundation; Araclon Biotech; BioClinica, Inc.; Biogen; Bristol-Myers Squibb Company; CereSpir, Inc.; Cogstate; Eisai Inc.; Elan Pharmaceuticals, Inc.; Eli Lilly and Company; EuroImmun; F. Hoffmann-La Roche Ltd and its affiliated company Genentech, Inc.; Fujirebio; GE Healthcare; IXICO Ltd.; Janssen Alzheimer Immunotherapy Research & Development, LLC.; Johnson & Johnson Pharmaceutical Research & Development LLC.; Lumosity; Lundbeck; Merck & Co., Inc.; Meso Scale Diagnostics, LLC.; NeuroRx Research; Neurotrack Technologies; Novartis Pharmaceuticals Corporation; Pfizer Inc.; Piramal Imaging; Servier; Takeda Pharmaceutical Company; and Transition Therapeutics. The Canadian Institutes of Health Research is providing funds to support ADNI clinical sites in Canada. Private sector contributions are facilitated by the Foundation for the National Institutes of Health (www.fnih.org). The grantee organization is the Northern California Institute for Research and Education, and the study is coordinated by the Alzheimer’s Therapeutic Research Institute at the University of Southern California. ADNI data are disseminated by the Laboratory for Neuro Imaging at the University of Southern California.

*Reference verification: 28 entries were resolved to specific PubMed records and checked field by field against NCBI efetch XML (title, first author, journal, year, volume, issue, page range or electronic article number, and DOI); entries 19 and 23 are monographs and entry 25 a physics-journal article not indexed in PubMed, transcribed from their original citations. The audit log is supplied as Supporting Information (REFERENCE_FIELD_AUDIT.log). No reference was generated without verification*.

## Supporting information

S1 Fig. Supporting validity and robustness checks. (Caption as listed above; file Figure_S1 provided separately in PNG, PDF and SVG.)

S1 Table. Per-night multifractal coupling features (mros_xml_features.csv; 3,928 rows x 53 columns).

S2 Table. Stage D quantitative synthesis (stageD_results.json; stageD_lambda_bootstrap.csv; stageD_power_lambda.csv; stageD_bias_reestimation.csv; stageD_spearman_brown.csv; stageD_montecarlo.csv; stageD_scorecard.csv).

S1 File. Complete analysis code (virtual tar reader, MF-DFA/DCCA, index construction, calibration, ADNI and MrOS association models, Firth module, quantitative synthesis and figure generation).

S2 File. Reproducibility and verification logs (final_manuscript_validation.log; REFERENCE_VERIFICATION_FINAL.log; REFERENCE_FIELD_AUDIT.log).

---

**Supplementary Figure S1.**
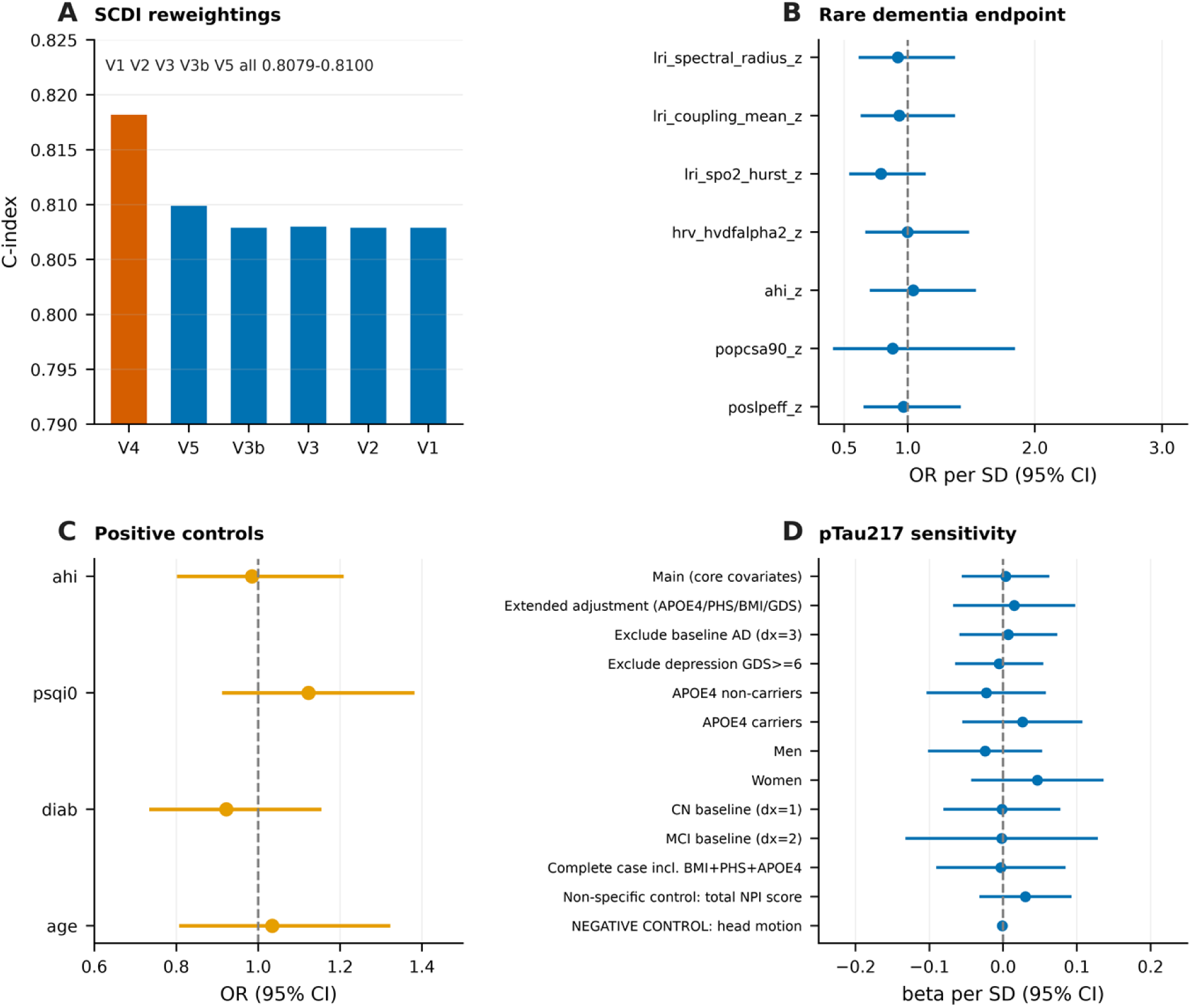
Supporting validity and robustness checks. (A) C-index of all five SCDI reweightings versus the post-hoc HRV long-range coupling fingerprint. (B) Firth-penalised logistic odds ratios for the 35-event self-reported dementia/AD endpoint in MrOS; all null after excluding baseline AD-medication users. (C) Positive-control checks for the constructed incident cardiovascular endpoint: age, diabetes, Pittsburgh Sleep Quality Index and AHI were all null, so this arm is classified as indeterminate rather than a failed replication. (D) Thirteen sensitivity/stratified analyses of the ADNI primary pTau217 estimate.

## Notes

### Competing Interest Statement

The authors have declared no competing interest.

### Author Declarations

he Sleep Heart Health Study, the Multi-Ethnic Study of Atherosclerosis, the MrOS Sleep Study and the Alzheimer's Disease Neuroimaging Initiative were approved by the institutional review boards of their participating institutions, and all participants provided written informed consent before enrolment. The present work is a secondary analysis of de-identified data released under the NSRR Data Access and Use Agreement and the ADNI Data Use Agreement. No new participants were recruited, no identifying information was accessed and no additional contact with participants took place consequently no separate ethical approval was required for this analysis. All procedures were conducted in accordance with the Declaration of Helsinki and with the data-use and publication policies of the source studies.

## References

1. Wang Z, Yin C, Cheng A, et al. Long-range interdependencies aware signal analysis for rapid and accurate obstructive sleep apnea prediction. Nat Commun. 2026;17(1):11024. doi:10.1038/s41467-026-76274-0.

2. Xie L, Kang H, Xu Q, et al. Sleep drives metabolite clearance from the adult brain. Science. 2013;342(6156):373–377.

3. Iliff JJ, Wang M, Liao Y, et al. A paravascular pathway facilitates CSF flow through the brain parenchyma and the clearance of interstitial solutes, including amyloid beta. Sci Transl Med. 2012;4(147):147ra111.

4. Ju YE, Lucey BP, Holtzman DM. Sleep and Alzheimer disease pathology - a bidirectional relationship. Nat Rev Neurol. 2014;10(2):115–119.

5. Yaffe K, Laffan AM, Harrison SL, et al. Sleep-disordered breathing, hypoxia, and risk of mild cognitive impairment and dementia in older women. JAMA. 2011;306(6):613–619.

6. Leng Y, McEvoy CT, Allen IE, et al. Association of sleep-disordered breathing with cognitive function and risk of cognitive impairment: a systematic review and meta-analysis. JAMA Neurol. 2017;74(10):1237–1245.

7. Himali JJ, Baril AA, Cavuoto MG, et al. Association between slow-wave sleep loss and incident dementia. JAMA Neurol. 2023;80(12):1326–1333.

8. Bubu OM, Kaur SS, Mbah AK, et al. Obstructive sleep apnea and hypertension with longitudinal amyloid-beta burden and cognitive changes. Am J Respir Crit Care Med. 2022;206(5):632–636.

9. Jack CR Jr, Bennett DA, Blennow K, et al. NIA-AA Research Framework: toward a biological definition of Alzheimer’s disease. Alzheimers Dement. 2018;14(4):535–562.

10. Palmqvist S, Janelidze S, Quiroz YT, et al. Discriminative accuracy of plasma phospho-tau217 for Alzheimer disease vs other neurodegenerative disorders. JAMA. 2020;324(8):772–781.

11. Quan SF, Howard BV, Iber C, et al. The Sleep Heart Health Study: design, rationale, and methods. Sleep. 1997;20(12):1077–1085.

12. Bild DE, Bluemke DA, Burke GL, et al. Multi-Ethnic Study of Atherosclerosis: objectives and design. Am J Epidemiol. 2002;156(9):871–881.

13. Zhang Y, Kim M, Prerau M, et al. The National Sleep Research Resource: making data findable, accessible, interoperable, reusable and promoting sleep science. Sleep. 2024;47(7):zsae088.

14. Mueller SG, Weiner MW, Thal LJ, et al. The Alzheimer’s Disease Neuroimaging Initiative. Neuroimaging Clin N Am. 2005;15(4):869–877.

15. Cummings JL, Mega M, Gray K, et al. The Neuropsychiatric Inventory: comprehensive assessment of psychopathology in dementia. Neurology. 1994;44(12):2308–2314.

16. Kaufer DI, Cummings JL, Ketchel P, et al. Validation of the NPI-Q, a brief clinical form of the Neuropsychiatric Inventory. J Neuropsychiatry Clin Neurosci. 2000;12(2):233–239.

17. Cespedes EM, Hu FB, Redline S, et al. Comparison of self-reported sleep duration with actigraphy: results from the Hispanic Community Health Study/Study of Latinos Sueno Ancillary Study. Am J Epidemiol. 2016;183(6):561–573.

18. Rezaie L, Fobian AD, McCall WV, Khazaie H. Paradoxical insomnia and subjective-objective sleep discrepancy: a review. Sleep Med Rev. 2018;40:196–202.

19. Carroll RJ, Ruppert D, Stefanski LA, Crainiceanu CM. Measurement Error in Nonlinear Models: A Modern Perspective. 2nd ed. Boca Raton: Chapman & Hall/CRC; 2006.

20. Alexeeff SE, Carroll RJ, Coull B. Spatial measurement error and correction by spatial SIMEX in linear regression models when using predicted air pollution exposures. Biostatistics. 2016;17(2):377–389.

21. Stöberl AS, Schwarz EI, Haile SR, et al. Night-to-night variability of obstructive sleep apnea. J Sleep Res. 2017;26(6):782–788.

22. Lechat B, Loffler KA, Reynolds AC, et al. High night-to-night variability in sleep apnea severity is associated with uncontrolled hypertension. NPJ Digit Med. 2023;6(1):57.

23. Nunnally JC, Bernstein IH. Psychometric Theory. 3rd ed. New York: McGraw-Hill; 1994.

24. Peng CK, Havlin S, Stanley HE, Goldberger AL. Quantification of scaling exponents and crossover phenomena in nonstationary heartbeat time series. Chaos. 1995;5(1):82–87.

25. Kantelhardt JW, Zschiegner SA, Koscielny-Bunde E, et al. Multifractal detrended fluctuation analysis of nonstationary time series. Physica A. 2002;316(1-4):87–114.

26. Podobnik B, Stanley HE. Detrended cross-correlation analysis: a new method for analyzing two nonstationary time series. Phys Rev Lett. 2008;100(8):084102.

27. Heinze G, Schemper M. A solution to the problem of separation in logistic regression. Stat Med. 2002;21(16):2409–2419.

28. Blackwell T, Yaffe K, Laffan A, et al. Associations of objectively and subjectively measured sleep quality with subsequent cognitive decline in older community-dwelling men. Sleep. 2014;37(4):655–663.

29. Cavaillès C, Yaffe K, Blackwell T, Buysse D. Multidimensional sleep health and long-term cognitive decline in community-dwelling older men. J Alzheimers Dis. 2023;96(1):65–71.

30. von Elm E, Altman DG, Egger M, et al. The Strengthening the Reporting of Observational Studies in Epidemiology (STROBE) statement: guidelines for reporting observational studies. J Clin Epidemiol. 2008;61(4):344–349.

31. Ioannidis JPA. Inverse publication reporting bias favouring null, negative results. BMJ Evid Based Med. 2024;29(1):6–9.

